# Association between post-infection COVID-19 vaccination and symptom severity of post COVID-19 condition among patients on Bonaire, Caribbean Netherlands: a retrospective cohort study

**DOI:** 10.1101/2023.06.20.23291649

**Authors:** D.S.F. Berry, T. Dalhuisen, G. Marchena, I. Tiemessen, E. Geubbels, L. Jaspers

**Author notes:** Corresponding author: Loes Jaspers. Joint first authorship (D.S.F. Berry, T. Dalhuisen).

## Abstract

**Objectives:** In this retrospective cohort study, we aimed to investigate symptom severity change following COVID-19 vaccination among post COVID-19 condition (PCC) patients on Bonaire.

**Methods:** Symptomatic cases who tested positive for SARS-CoV-2 between the start of the pandemic and 1 October 2021, were unrecovered on the interview day and unvaccinated prior to infection were identified from the national case registry. Patients were interviewed by telephone between 15 November and 4 December 2021 about sociodemographic factors, pre-pandemic health, COVID-19 symptoms and vaccination status. We compared symptom severity change between the acute and post-acute disease phase (>4 weeks after disease onset) of 14 symptoms on a five-point Likert scale for 36 PCC patients having received at least one dose of the BNT162 (BioNTech/Pfizer) vaccine and 11 patients who remained unvaccinated, using separate multiple linear regression models.

**Results:** Most common post-acute symptoms included fatigue (81%), reduced physical endurance (79%), and reduced muscle strength (64%). Post-infection vaccination was significantly associated with reduced severity of heart palpitations, after adjusting for acute phase severity and duration of illness (β 0.60, 95% CI 0.18-1.02). We did not find a statistically significant association with symptom severity change for other, more prevalent symptoms.

**Conclusions:** Larger prospective studies are needed to confirm our observation in a small study population that post-infection COVID-19 vaccination was associated with reduced severity of heart palpitations among those with this symptom self-attributed to SARS-CoV-2 infection.

## Introduction

As of March 2020, health systems became severely challenged by the global outbreak of SARS-CoV-2; the agent responsible for the coronavirus disease 19 (COVID-19). Concurrently, countries have been reporting increasing evidence of an emerging syndrome of prolonged disease among COVID-19 patients, where up to one in three (34%, 95% CI 26-49%) cases infected with SARS-CoV-2 may continue to experience symptoms longer than 30 days after disease onset and 32% of cases at 90 days post-infection (95% CI 14-57%) [1]. This condition is commonly referred to as ‘long-COVID’, ‘post-acute COVID-19’, or ‘post COVID-19 condition (PCC)’. Fatigue, headache, and brain fog are commonly reported post-acute symptoms, though shortness of breath, reduced muscle strength, anosmia, cognitive impairment, and many others have been reported as well [1–3].

Even though COVID-19 vaccines have been proven effective in preventing severe COVID-19 disease, much remains unknown about their effect on PCC [3]. Similar to post-infectious syndromes such as those following Borreliosis, Chikungunya, Ebola, and MERS, PCC is characterized by immunological dysregulation and the presence of long-term sequelae, presented through a variety of symptoms across organ systems [2, 3]. Biological pathways underlying PCC remain unclear, though it is hypothesized that reactivation of the Epstein-Barr virus, persistent reservoirs of SARS-CoV-2, a dysregulated or auto-immune response, platelet hyperactivation and microclot formation or cell dysmetabolism may play a role [2–4]. Using COVID-19 vaccination as a therapeutic intervention may aid cellular metabolism to eliminate viral residue and restore the antiviral immune response, thereby alleviating symptom severity [2, 3].

Studies on the effect of vaccination given after acute SARS-CoV-2 infection in unvaccinated PCC patients report mixed effects on post-acute symptoms, suggesting either improvement, worsening, or no change in symptomology after vaccination [3, 5, 6]. A recent systematic review including 16 observational studies from five Western countries, among which the Netherlands, showed receiving either one (OR 0.22-1.03) or two doses (two doses OR 0.25-1.02) of COVID-19 vaccination prior to SARS-CoV-2 infection significantly reduced the incidence of PCC [7].

Bonaire is an island in the Dutch Caribbean and has 21,745 permanent residents, details can be found in the Central Bureau of Statistics (CBS) database [8]. Between early March 2020 and 1 October 2021, 2,036 SARS-CoV-2 infections were confirmed on Bonaire, having led to 57 hospital admissions and 19 COVID-19 related deaths by that time [9]. To date, it remains unknown what the prevalence of PCC is on Bonaire and to what extent vaccination impacts the severity of post-acute COVID-19 symptoms. In this study, we aim to examine the association between vaccination status after SARS-CoV-2 infection, using the Pfizer vaccine, and change in self-reported symptom severity among unrecovered PCC patients on Bonaire.

## Methods

### Study design, data collection, and study population

This sub-study is part of an overarching retrospective cohort study looking into the prevalence and symptomatology of PCC on Bonaire [Berry et al. (2023), ‘unpublished observations’]. Data was collected through telephone interviews between 15 November and 4 December 2021. We defined a PCC patient as “an individual with a laboratory confirmed SARS-CoV-2 positive test result, for whom at least one symptom self-attributed to the experienced SARS-CoV-2 infection lasted longer than four weeks” [5]. The telephone survey included questions on comorbidity and pre-existing conditions, hospitalization during acute COVID-19 (defined as symptoms occurring within 4 weeks after disease onset), symptom prevalence and severity during the acute and post-acute phase, date(s) and type of COVID-19 vaccination, and demographic and lifestyle data. For the overarching retrospective cohort study, a power analysis was done using an estimated prevalence of PCC of 20% and expected response rate of 70%. A description of the full methodology and survey can be found in [Berry et al. (2023), ‘unpublished observations’].

For this sub-study, we used self-reported data of PCC patients who were unvaccinated prior to SARS-CoV-2 infection and who were unrecovered at the time of interview (**Figure 1**). We could not use data for PCC patients who were already recovered at the time of interview, because our data did not have sufficient temporal resolution to determine when exactly these patients recovered relative to their COVID-19 vaccination date(s). Additionally, in our opinion, using data of unrecovered PCC patients also minimized recall bias regarding symptom severity.

**Figure 1:**
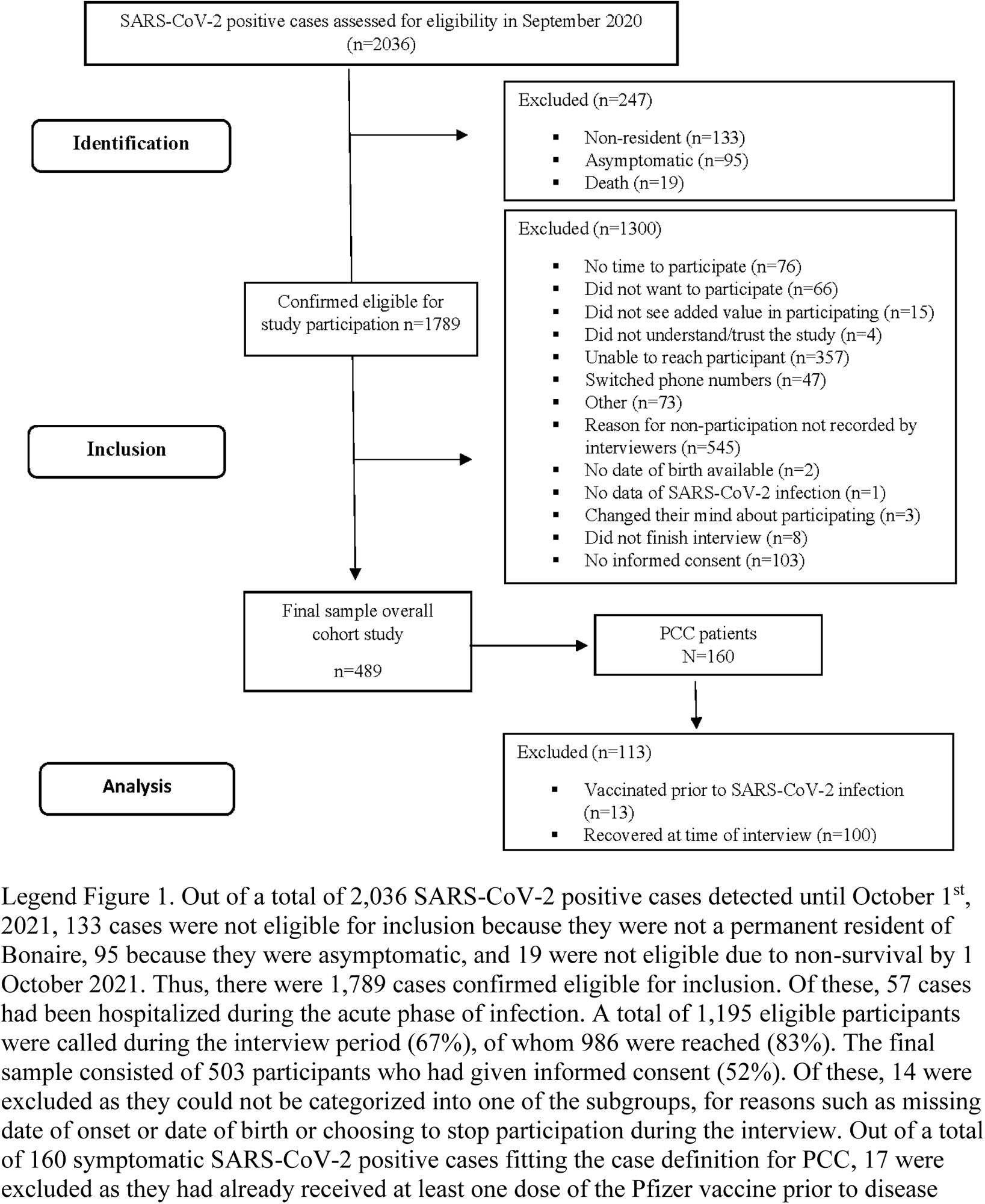
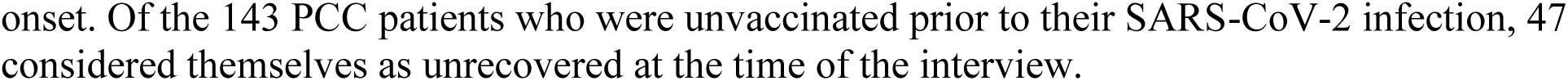
STROBE flowchart describing participant selection for this sub-study examining the association between post-infection COVID-19 vaccination and symptom severity change of PCC patients on Bonaire, Caribbean Netherlands.

### Exposure and outcome variables

On Bonaire, the COVID-19 vaccination campaign started mid-February 2021 using the BNT162b2 (BioNTech/Pfizer) vaccine, further referred to as Pfizer [9]. Dates of first COVID-19 vaccination and onset of disease were used to determine vaccination status of PCC patients before and after their SARS-CoV-2 infection. We used a cutoff of at least 14 days between the onset date of acute COVID-19 and date of first COVID-19 vaccination to limit the possibility of self-reported symptoms being attributable to COVID-19 vaccine side effects rather than attributable to the experienced SARS-CoV-2 infection. We did not differentiate subgroups by number of doses received due to the study’s small sample size and international heterogeneity in the definitions of COVID-19 vaccination status, but instead considered those vaccinated as having received at least one dose of the Pfizer vaccine at least 8 weeks after SARS-CoV-2 infection, in accordance with the recommendations on Bonaire at the time of the study [10].

The primary outcome was change in severity between the acute and post-acute phase (defined as symptoms persisting for more than four weeks after disease onset) of the following 14 COVID-19 symptoms: headache, heart palpitations, shortness of breath, cough, chest pain, worsened endurance, reduced muscle strength, muscle ache, loss of smell, loss of taste, loss of appetite, concentration problems, sleep changes, and fatigue. Using an ordinal five-point Likert scale, participants reported the severity of each symptom during the acute and post-acute phase, with 0=absent, 1=mild, 2=moderate, 3=severe, and 4=very severe. For each PCC patient and symptom, a change in severity score was calculated by subtracting the severity score of post-acute symptoms from the severity score during acute COVID-19. A positive change in severity score therefore indicates an improvement of symptom severity score in the post-acute phase compared to the acute phase. Our outcome of interest was to determine whether a positive or negative change in symptom severity score occurred among patients who received post-infection vaccination, whilst controlling for several factors.

### Covariates

Age and BMI were included as continuous variables. Hospital admission status during acute COVID-19 was coded as a binary variable (yes/no). We did not differentiate between admission to ICU or general ward and did not include length of hospital stay as a covariate due to the small sample of hospitalized patients in our study. PCC patients were asked whether they smoked cigarettes or drank alcohol prior to the SARS-CoV-2 pandemic (yes/no) and whether they had underlying comorbidities, which we recoded into a binary variable (yes/no). This included 13 comorbidities; for a full list see **Supplementary Table S1**. Respondents were asked to score their pre-pandemic health on a scale of 0 to 100, with 0 being the worst imaginable health and 100 the best imaginable health [11]. The disease onset date was considered as the first date of reporting any COVID-19 symptoms and duration of illness was calculated as the number of weeks between the interview date and the disease onset date.

### Statistical analyses

Differences in sex, hospitalization status, smoking status, alcohol use, and comorbidity by vaccination status were assessed using Fisher’s exact test. Differences in duration of illness, prepandemic health score, body mass index (BMI), and age between the two groups were assessed using the Kruskal-Wallis test. There were no records with missing data for the covariates included in the analyses. Univariable linear regression models were fitted to estimate coefficients and their 95% confidence intervals (CIs) for the association between vaccination status and change in severity score. A separate model was fitted for each symptom as we hypothesized that disparate biological pathways underlie each symptom [12, 13]. Next, multiple linear regression models were fitted to adjust for confounding variables.

Local clinicians identified a set of potentially confounding variables, i.e. BMI, age, sex, symptom severity during acute COVID-19, duration of illness, pre-pandemic health score, smoking status, alcohol use, having been hospitalized with acute COVID-19, and having at least one underlying comorbidity (**Supplementary Table S1**). Using this list of potential confounders, we fitted multiple linear regression models including vaccination status after infection and one potential confounding variable for each symptom. If the effect estimate change of vaccination status was larger than 10% compared to the univariable model, we included this confounder in the next model building step.

Next, we fitted multiple linear regression using the confounding variables identified in the previous step. Starting with the variable that had the largest vaccination status effect estimate change, we iteratively added one variable to the model and assessed the model’s AIC. If adding a confounding variable improved model fit (i.e. rule of thumb AIC decreased by more than 2 points) then the variable was added to the model. The model with the lowest AIC was chosen as the final adjusted model and preference was given to sparser models if the AIC did not decrease by more than 2 points. No more than four confounding variables were included in each model. *P* values < 0.05 were considered statistically significant. All analyses were carried out in R version 4.1.3 (R Foundation for Statistical Computing).

### Potential bias

We compared unrecovered patients with recovered patients on self-reported pre-pandemic health score, age, and BMI to investigate whether there may have been selection bias due to differences in recovery speed between patients who were vaccinated post-infection and those who remained unvaccinated, which could have resulted in different recovery potential between both groups.

### Sensitivity analysis

The median disease onset date was over one month earlier for vaccinated patients than for patients who remained unvaccinated. Consequently, the duration of illness at the time of the interview was shorter among patients who remained unvaccinated (32 weeks), as compared to those who subsequently had gotten vaccinated (37 weeks) (Kruskal-Wallis, p < 0.05). This suggests patients who remained unvaccinated had less time to recover compared to vaccinated patients. To account for this difference, we performed a sensitivity analysis to assess the effect of adding duration of illness as a covariate in our models. First, we excluded duration of illness from the best fitting models for the symptoms that included duration of illness as a covariate. Second, we rebuilt our models not including duration of illness in our covariate selection steps which changed the covariates included in the best final adjusted models. Last, we added duration of illness to these new adjusted final models and assessed changes in effect size estimates and model conclusions (**Supplementary Table S2**).

## Results

### Population characteristics

Out of a total of 160 symptomatic SARS-CoV-2 positive cases fitting the case definition for PCC, 17 were excluded as they had already received at least one dose of the Pfizer vaccine prior to disease onset. Of the 143 patients who were unvaccinated prior to their SARS-CoV-2 infection, 47 (33%) considered themselves as unrecovered at the time of the interview. Therefore, our final sample included 47 PCC patients (**Figure 1**).

The median age was 47 years (range 14 – 89 years old). Most (79%) patients in our sample were female (79%) and overweight (median BMI 31.2). Eleven (23%) patients reported having at least one pre-existing condition and 7 (15%) patients were admitted to the hospital during acute COVID-19. The majority of patients drank alcohol prior to the pandemic (60%), and two patients had a history of smoking (4%).

After infection, 11 (23%) patients remained unvaccinated, whereas 36 (76%) patients were vaccinated with at least one dose of the Pfizer vaccine. Patients who remained unvaccinated after infection were older than vaccinated patients (median 51 and 44 years old respectively) and had a lower BMI (median 30.7 and 31.9 respectively). For all patients, the self-assessment of their pre-pandemic health state was on the high end of the EQ-VAS scale (median scores 84 and 85 out of 100). The median disease onset date of vaccinated patients was about one month earlier than of those who remained unvaccinated, 12 March 2021 and 15 April 2021 respectively. The median time between dates of disease onset and first vaccination was 8.3 weeks, which suggests patients chose to get vaccinated soon after they were eligible for post-infection vaccination (which was 8 weeks after infection at the time of the study). Median duration of illness among vaccinated PCC patients was 37 weeks and 32 weeks among unvaccinated PCC patients (**Table 1**).

**Table 1:** Baseline characteristics of the study population by post-infection vaccination status^1^.

|  | Vaccination status of unrecovered PCC patients after SARS-CoV-2 infection |  |  |
| --- | --- | --- | --- |
|  | Vaccinated (n = 36) | Unvaccinated (n = 11) | P-value |
| Median age (IQR) (years) | 44 (19.2) | 51 (16.5) | 0.36<sup>A</sup> |
| Median BMI (IQR) | 30.7 (8.8) | 31.9 (14.8) | 0.93<sup>A</sup> |
| Sex |  |  | 1.00<sup>B</sup> |
| Male | 8 (22%) | 2 (18%) |  |
| Female | 28 (78%) | 9 (82%) |  |
| Median prepandemic health score (IQR) (0-100) | 83.5 (13.5) | 85.0 (13.5) | 0.16<sup>A</sup> |
| Prepandemic smoker |  |  | 1.00<sup>B</sup> |
| Yes | 2 (6%) | 0 (0%) |  |
| No | 34 (94%) | 11 (100%) |  |
| Prepandemic alcohol use |  |  | 0.74<sup>B</sup> |
| Yes | 22 (61%) | 6 (55%) |  |
| No | 14 (39%) | 5 (45%) |  |
| Prepandemic comorbidity |  |  | 0.70<sup>B</sup> |
| Yes | 10 (28%) | 2 (18%) |  |
| No | 26 (72 %) | 9 (82%) |  |
| Hospital admission during acute COVID-19 |  |  | 0.33<sup>B</sup> |
| Yes | 4 (11%) | 3 (27%) |  |
| No | 32 (89%) | 8 (73%) |  |
| Median time between dates of disease onset and first vaccination (IQR) (weeks) | 8.3 (11) | - | - |
| Median duration of illness (IQR) (weeks) | 36.6 (3.4) | 32.1 (18.0) | **<0.001<sup>A</sup>** |
| Median acute phase severity score (IQR) |  |  |  |
| Chest pain | 0 (2.0) | 1.0 (2.0) | 0.95<sup>A</sup> |
| Concentration problems | 1.0 (2.0) | 2.0 (1.50) | 0.09<sup>A</sup> |
| Cough | 2.0 (3.0) | 1.0 (1.0) | 0.82<sup>A</sup> |
| Fatigue | 3.0 (2.25) | 2.0 (1.0) | 0.63<sup>A</sup> |
| Headache | 2.0 (1.0) | 2.0 (1.50) | 0.23<sup>A</sup> |
| Heart palpitations | 0.0 (2.0) | 0 (0.50) | 0.37<sup>A</sup> |
| Loss of appetite | 2.0 (3.0) | 2.0 (3.0) | 0.66<sup>A</sup> |
| Reduced muscle strength | 2.0 (2.0) | 2.0 (2.0) | 0.86<sup>A</sup> |
| Loss of sense of smell | 1.0 (3.0) | 2.0 (3.0) | 0.77<sup>A</sup> |
| Loss of sense of taste | 1.5 (3.0) | 3.0 (2.5) | 0.24<sup>A</sup> |
| Muscle ache | 2.0 (2.0) | 2.0 (2.5) | 0.84<sup>A</sup> |
| Reduced physical endurance | 2.5 (2.0) | 2.0 (1.5) | 0.83<sup>A</sup> |
| Shortness of breath | 2.0 (3.0) | 2.0 (2.0) | 0.96<sup>A</sup> |
| Sleeping problems | 2.0 (3.0) | 2.0 (2.5) | 0.86<sup>A</sup> |

<sup>1</sup> Interquartile range (IQR); Body mass index (BMI); Post COVID-19 condition (PCC).
<sup>A</sup> Wilcoxon test. <sup>B</sup> Fisher's exact test.

### Symptomatology

#### Acute COVID-19

Common symptoms during the first four weeks of infection included fatigue (94%), reduced muscle strength (87%), and reduced physical endurance (87%). Patients who remained unvaccinated more often reported experiencing concentration problems (91% vs. 53%), reduced physical endurance (100% vs. 83%), cough (82% vs. 67%), loss of sense of taste (73% vs. 61%), sleeping problems (73% vs. 64%), chest pain (55% vs. 47%), fatigue (100% vs. 92%), shortness of breath (73% vs. 67%), and reduced muscle strength (91% vs. 86%) than vaccinated patients (**Table 2**). For both groups, heart palpitations (36% and 26%) and chest pain (47% and 55%) were the least prevalent symptoms.

**Table 2:** Number and percentage of unrecovered PCC patients who self-report presence of COVID-19 symptoms during acute COVID-19 and the post-acute phase of infection, stratified by post-infection vaccination status.

|  | **Vaccinated**<br>**(n = 36)** |  | **Remained unvaccinated**<br>**(n = 11)** |  | **Overall**<br>**(n = 47)** |  |
| --- | --- | --- | --- | --- | --- | --- |
| **Symptom** | **Acute**<br>**phase** | **Post-**<br>**acute**<br>**phase** | **Acute**<br>**phase** | **Post-acute**<br>**phase** | **Acute**<br>**phase** | **Post-acute**<br>**phase** |
| Chest pain | 17 (47%) | 12 (33%) | 6 (55%) | 5 (45%) | 23 (49%) | 17 (36%) |
| Concentration problems | 19 (53%) | 20 (56%) | 10 (91%) | 8 (73%) | 29 (62%) | 28 (60%) |
| Cough | 24 (67%) | 18 (50%) | 9 (82%) | 5 (45%) | 33 (70%) | 23 (49%) |
| Fatigue | 33 (92%) | 29 (81%) | 11 (100%) | 9 (82%) | 44 (94%) | 38 (81%) |
| Headache | 31 (86%) | 15 (42%) | 9 (82%) | 8 (73%) | 40 (85%) | 23 (49%) |
| Heart palpitations | 13 (36%) | 10 (28%) | 3 (27%) | 4 (36%) | 16 (34%) | 14 (30%) |
| Loss of appetite | 25 (69%) | 9 (25%) | 6 (55%) | 4 (36%) | 31 (66%) | 13 (28%) |
| Reduced muscle strength | 31 (86%) | 21 (58%) | 10 (91%) | 9 (73%) | 41 (87%) | 30 (64%) |
| Loss of sense of smell | 20 (56%) | 12 (33%) | 6 (55%) | 6 (55%) | 26 (55%) | 18 (38%) |
| Loss of sense of taste | 22 (61%) | 12 (33%) | 8 (73%) | 6 (55%) | 30 (64%) | 18 (38%) |
| Muscle ache | 29 (81%) | 20 (56%) | 8 (73%) | 9 (82%) | 37 (79%) | 29 (62%) |
| Reduced physical endurance | 30 (83%) | 28 (78%) | 11 (100%) | 9 (82%) | 41 (87%) | 37 (79%) |
| Shortness of breath | 24 (67%) | 21 (58%) | 8 (73%) | 8 (73%) | 32 (86%) | 29 (62%) |
| Sleeping problems | 23 (64%) | 18 (50%) | 8 (73%) | 6 (55%) | 31 (66%) | 24 (51%) |

#### Post-acute phase

Fatigue (81% vs. 82%), reduced physical endurance (79% vs. 82%), and reduced muscle strength (64% vs. 82%) were the most prevalent symptoms in both groups. Apart from cough, all symptoms were more prevalent among patients who remained unvaccinated than among vaccinated PCC patients. For vaccinated patients, all symptoms were less prevalent in the post-acute phase compared to during acute COVID-19, with exception of concentration problems. The largest decrease in prevalence was observed for loss of appetite (69% to 25%) and headache (86% to 42%), while the prevalence of heart palpitations (36% to 28%), reduced physical endurance (83% to 78%), and shortness of breath (67% to 58%) remained stable between the two phases. Among patients who remained unvaccinated, the most persistent symptoms included headache (82% to 73%) and chest pain (55% to 45%), and the prevalence of heart palpitations and muscle ache increased slightly (27% to 36% and 73% to 82% respectively).

### Severity

The full range of symptom severity scores (0-4) was reported by patients during acute COVID-19 and during the post-acute phase (**Sup Figures 1 & 2**). A frequency table with the severity scores in the acute and post-acute phase can be found in **Supplementary Table S3**. Median severity scores tended to be lower in the post-acute phase compared to during acute COVID-19 and more symptoms were reported as absent compared to the acute phase.

### Change in symptom severity

Relative to patients who remained unvaccinated after infection, vaccination after infection was associated with an improvement in the severity of headache and heart palpitations, but not for other symptoms (**Figures 2** and **3**, **Supplementary Tables 4** and **5**).

**Figure 2:**
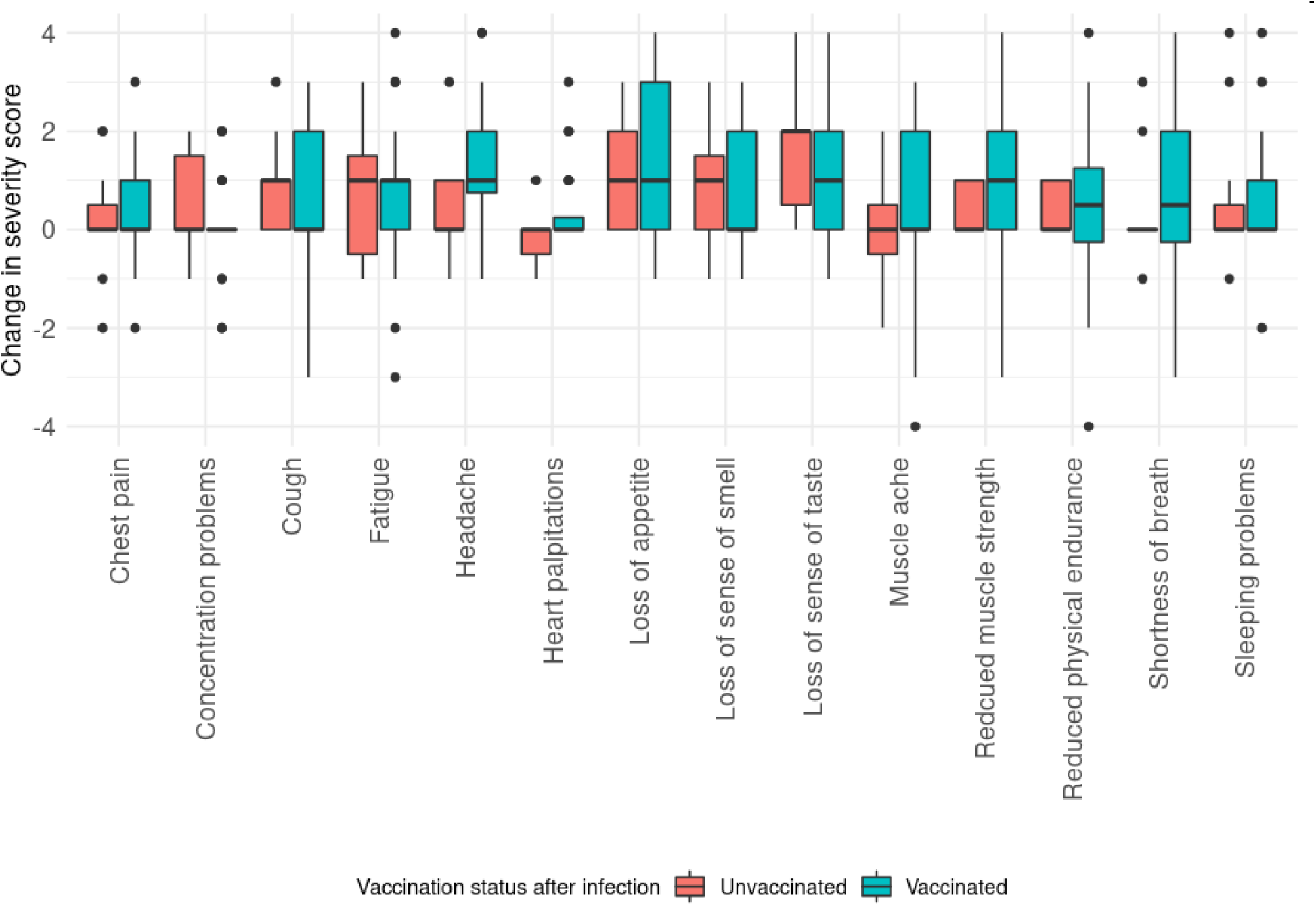
Boxplot showing the change in severity score between the post-acute phase and acute phase of infection per symptom for all 14 symptoms and PCC patients (n = 47) stratified by vaccination status after infection. A positive value for the change in severity score indicates an improvement of symptom severity subsequent to vaccination. The change in severity score was calculated by subtracting the acute COVID-19 severity score from the post-acute phase severity score. Both scores range from 0 (did not experience the symptom) to 4 (very severe).

**Figure 3:**
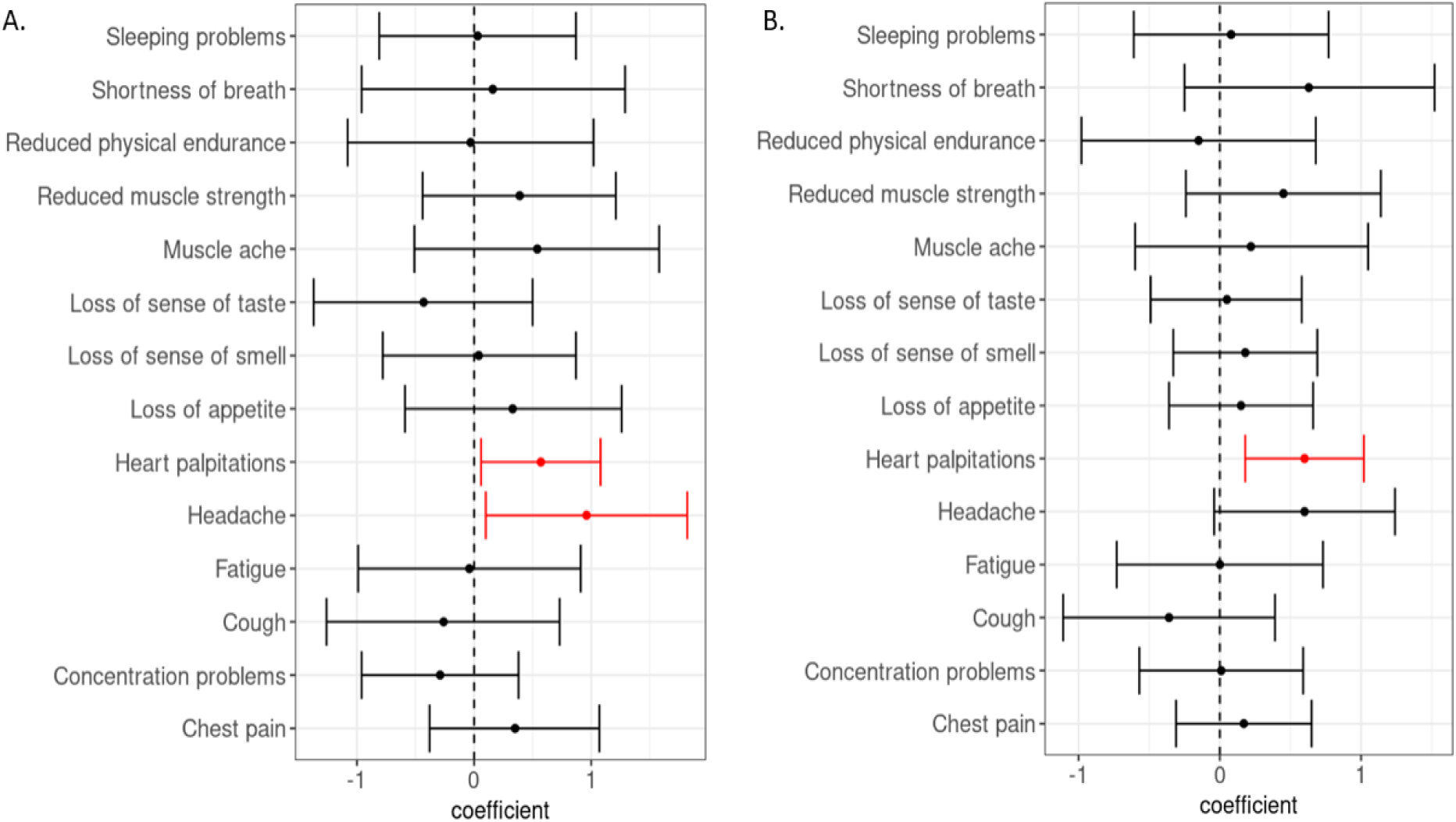
Association between vaccination status and change of symptom severity for 14 different symptoms with 95% CI. Reference category for all models is being unvaccinated and coefficient for being vaccinated is shown. Statistically significant coefficients are shown in red. A. Unadjusted multivariable models B. Multivariable models. For an overview of full model specification see Supplementary Table S3.

#### Headache

Among all patients reporting persisting headache (n = 23, 49%), the self-reported improvement was nearly one point stronger (β 0.96, p = 0.03, 95% CI 0.10, 1.82) on a scale of 0 to 4 for those who received post-infection vaccination (n = 15, 65%), as compared to patients with persisting headache who remained unvaccinated (n = 8, 35%). The decrease in severity of headache was no longer significant after adjusting for acute COVID-19 severity score (β 0.60, p = 0.06, 95% CI – 0.04, 1.24) (**Figures 2** and **3**).

The prevalence of headache among vaccinated PCC patients decreased from 86% to 42% between acute COVID-19 and the post-acute phase. None of the vaccinated PCC patients were experiencing very severe post-acute headache, and nearly two-thirds less (n = 6, 13%) were still experiencing severe post-acute headache. Among the 11 PCC patients who remained unvaccinated, headache was one of the most persistent symptoms, the prevalence reduced slightly from 82% to 73% in the post-acute phase. None of the unvaccinated PCC patients reported very severe or severe headache in the post-acute phase, whereas 33% reported severe headache in the acute phase, indicating severity decreased moderately over time (**Supplementary Table S3**).

#### Heart palpitations

The severity of heart palpitations reduced by over half a point (β = 0.57, p = 0.03, 95% CI 0.06, 1.08) among vaccinated patients (n = 10, 28%), as compared to those who remained unvaccinated (n = 4, 36%) (**Figures 2 and 3**). The effect of vaccination after infection on severity of heart palpitations remained significant after adjusting for acute COVID-19 severity and duration of illness (β=0.60, p=0.01, 95% CI 0.18, 1.02). Among vaccinated patients, the prevalence of heart palpitations reduced minimally between the acute and post-acute phase (36% to 28%), and heart palpitations were most often experienced as moderate (46%). The change in severity score was less apparent among PCC patients who remained unvaccinated, where the prevalence of heart palpitations increased slightly from 27% to 36%, and severity remained mild to moderate over time (**Supplementary Table S3**).

### Additional analyses

Excluding duration of illness from the best fitting model decreased the effect size for vaccination status and heart palpitations (0.60 to 0.37) and changed the p-value from 0.01 to 0.06 (**Supplementary Table S2**). Excluding duration of illness also decreased the effect size for vaccination status on shortness of breath (0.63 to 0.19), and its p-value increased from 0.16 to 0.63. For the second set of models, we found similar results, i.e. excluding duration of illness lead to a smaller effect size of vaccination status (**Supplementary Table S2**).

Furthermore, there was no evidence suggesting that change in symptom severity decreased as time between onset of disease and first vaccination increased, as evidenced by a low correlation coefficient (r = -0.09, p = 0.06, Pearson Correlation). This implies that those who had gotten vaccinated sooner after infection did not have a differential symptom severity change. Lastly, there was no statistically significant difference between recovered and unrecovered patients for median pre-pandemic health score (80 vs. 82 respectively, p = 0.43), age (41 vs. 44 years respectively, p = 0.21), and BMI (30.4 vs. 30.1, p = 0.20). Additionally, there was no difference in the proportion of recovered (73%) and unrecovered (77%) patients who received post-infection vaccination (p = 0.92).

## Discussion

Our results show a significantly larger improvement in the severity of post-acute heart palpitations for PCC patients who received post-infection vaccination compared to those who remained unvaccinated. Additionally, we found a reduced prevalence of most COVID-19 symptoms in the post-acute phase. This improvement appeared larger among vaccinated patients when compared to patients who remained unvaccinated, though none of the associations for other symptoms than heart palpitations were statistically significant. Thirty per cent of patients in our sample experienced post-acute heart palpitations, with a small difference in prevalence among vaccinated (28%) and unvaccinated patients (36%). In our sample, the overall prevalence of heart palpations was low compared to symptoms such as fatigue (94%) and reduced muscle strength (87%), for which we did not observe a significant change in symptom severity after COVID-19 vaccination.

Beyond cardiac injuries as myocardial inflammation or myocarditis [13–15], few studies have reported on other, less prevalent cardiovascular impact in hospitalized and non-hospitalized PCC patients. A study of PCC symptomatology at three months after infection among a cohort of European Dutch cases found the presence of palpitations or tachycardia in cases was associated with possible PCC, though they did not find the prevalence differed by vaccinated status [16]. In a Chinese cohort study [17], 9% of patients experienced tachycardia at six months following infection, and a study of 138 patients hospitalized in Wuhan found arrythmia’s were observed more frequently among ICU patients compared to non-ICU patients [18].

Though post-acute heart palpitations are not among the most common PCC symptoms in our study or in recent literature [1-3, 12, 14], the severity of these complaints may still interfere with PCC patients’ return to normal life as much as other, more common PCC symptoms [19]. Heart palpitations may also occur in PCC patients as a symptom of psychological distress such as anxiety, depression, or stress, or as a symptom of thyroid dysregulation [20] rather than a symptom of cardiovascular disease (CVD). Our findings shed light on the relevance of conducting more research into the potential biological mechanisms underlying dysrhythmias following SARS-CoV-2 infection [21], and how post-infection vaccination may play a role in alleviating the burden of lingering symptoms following COVID-19 disease.

### Methodological considerations

Our model adjustments for acute COVID-19 severity and duration of illness lead to our estimates being conservative. Acute COVID-19 severity was included as a potential confounder in all the multivariable models. Since the maximum possible change in severity score is dependent on the severity score in the acute phase and because we hypothesized that patients with a more severe acute infection would have been more likely to seek subsequent COVID-19 vaccination due to a local media coverage by a well-known radio-host, we chose to include the acute phase COVID-19 severity score of the respective symptom as a potential confounder. [22] Similar research suggests correcting for number of symptoms during acute COVID-19 may be an alternative indicator as opposed to correcting for acute COVID-19 severity [21].

Similarly, we opted to include duration of illness in the models, hypothesizing that those with a longer duration of illness are more prone to get vaccinated, as well as more prone to experience a change in severity of symptoms over time. Lastly, our analyses only assessed a numerical change in symptom severity score and care should be taken with interpreting these findings as clinical improvement. Given the ordinal nature of the symptom severity scores, a numeric change of 1 point in our findings may not scale evenly with a 1 point change between symptom levels on the five-point Likert scale.

### Strengths, limitations, and directions for future research

To our knowledge, this is the first study of PCC in the Caribbean setting and the first to provide insights into the symptomatology and severity of PCC among patients vaccinated after SARS-SoV-2 infection in the (Dutch) Caribbean context to date. Therefore, the results could be of interest to the broader Caribbean region. A second strong point of this study is that we had no missing data and therefore did not have to impute data or exclude cases. Another strength is that our sample included both hospitalized and non-hospitalized patients, and therefore the results of this study are relevant to the general population.

We used a patient-reported outcome (PRO) to assess symptom severity in each phase and did not add interpretation of these scores by a clinician. We believed using a PRO to determine symptom severity instead of a measure of severity rated by a GP would gain more valuable insights, as including patients’ experience of PCC would provide a more holistic interpretation of the hypothesized clinical benefit of post-infection vaccination [23]. By focusing on the sample of unrecovered PCC patients we attempted to reduce recall bias, as this group of patients was still experiencing symptoms at the time of interview. We believe asking recovered PCC patients to report the date when they first noticed remission of symptoms [24] would likely introduce recall bias, as PCC patients with a disease onset date from early 2021 may have been less likely to accurately report the date of recovery during the time of interview (15 November to 4 December 2021). We did not find a statistically significant difference between recovered and unrecovered PCC patients for median prepandemic health score, age, BMI, and the proportion of recovered PCC patients and unrecovered PCC patients who received post-infection vaccination, and therefore concluded selection bias related to recovery speed would not be likely in our sample of unrecovered PCC patients.

An important limitation is our small sample size, which hampers power to find statistically significant associations between post-infection vaccination and the severity of more prevalent symptoms. Although we adjusted in our analyses for known characteristics, residual confounding may exist due to unmeasured confounding variables, e.g. those associated with getting vaccinated and with post-acute symptoms. Also, we collected outcome data at one point in time, at different intervals since infection (and vaccination, for those applicable), limiting us in comparing severity scores of PCC symptoms at multiple moments after initial infection at an individual level. Lastly, we opted for multiple testing, hypothesizing that the causal biological pathways may differ per COVID-19 symptom [12, 13, 19]. We acknowledge that this could increase the chance of incidental findings due to our small sample size. Results from our study therefore would need to be reproduced by larger, preferably prospective and controlled studies, and be supported with insights into biological pathways, before any causal inference can be drawn on the negative association between COVID-19 vaccination status and prevalence of heart palpitations in the post-acute phase.

## Data Availability

All data produced in the present study are available upon reasonable request to the authors

## Acknowledgements

We would like to thank Alicia Krijgsman and Karin Cox (Public Health Department Bonaire), members of CBS Bonaire, and interviewers of the Tempo team on Bonaire for their contributions in the design and preparation of this study, data collection, and logistical assistance with implementing this study on Bonaire. We would like to thank Caroline van den Ende (RIVM, Cib, EPI) for advising on the latest publications regarding PCC.

## Authors contributions

DSF Berry: Conceptualization, methodology, resources, visualization, writing – original draft; submitting author.

T Dalhuisen: Data curation, formal analysis, methodology, software, visualization, writing – original draft.

G Marchena: Conceptualization, investigation, project administration. I Tiemessen: Investigation, data curation.

E Geubbels: Methodology, supervision, visualization, writing – original draft.

L Jaspers: Conceptualization, data curation, funding acquisition, investigation, methodology, project administration, supervision, visualization, writing – original draft, corresponding author.

## Data Availability Statement

Supplementary Material is available on request.

## Supplementary material

**Supplementary Figure 1:**
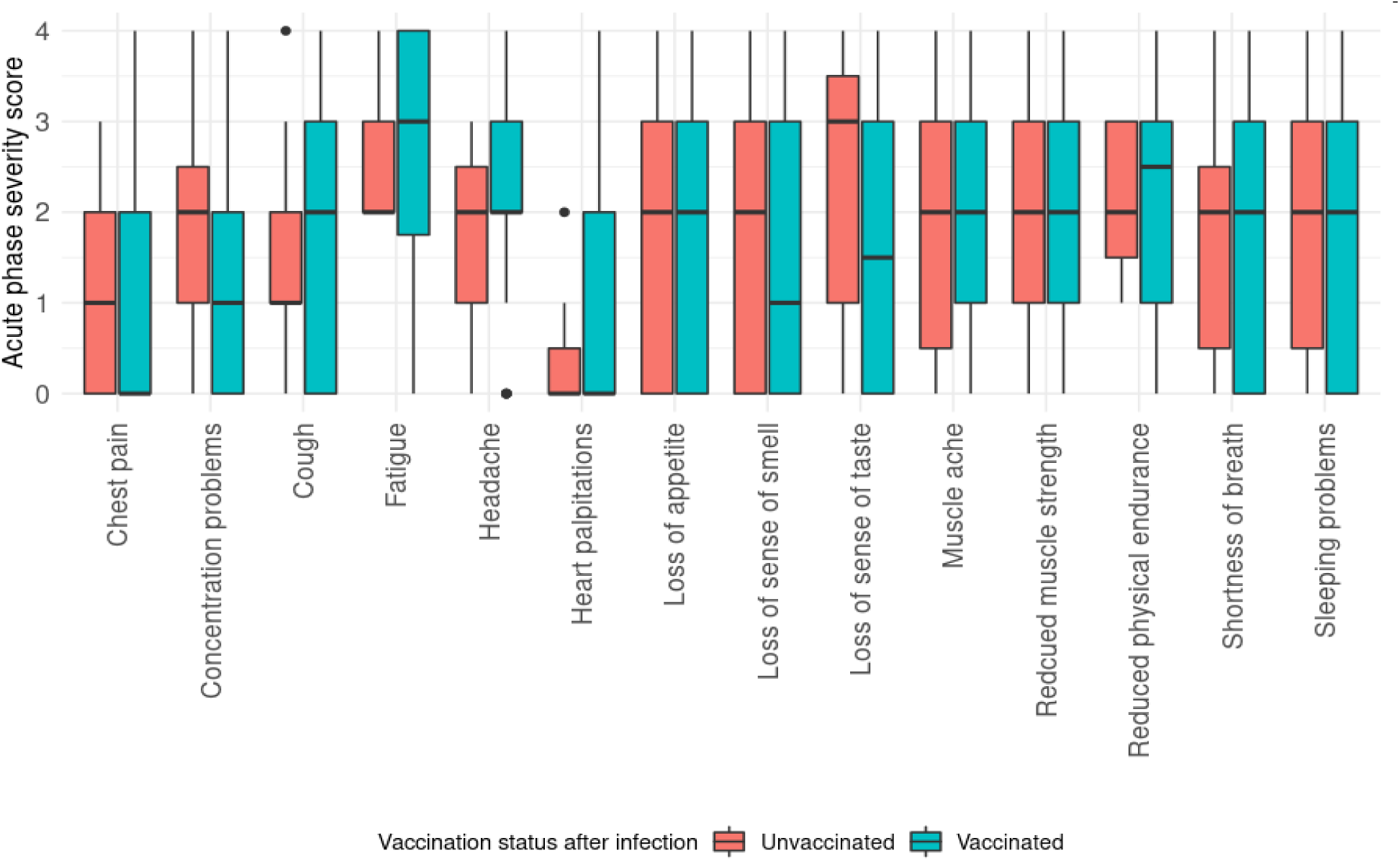
Boxplots showing acute COVID-19 severity scores for all 14 symptoms and PCC patients (n = 47) stratified by vaccination status after infection. Symptom severity scores range from 0 (did not experience the symptom) to 4 (very severe).

**Supplementary Figure 2:**
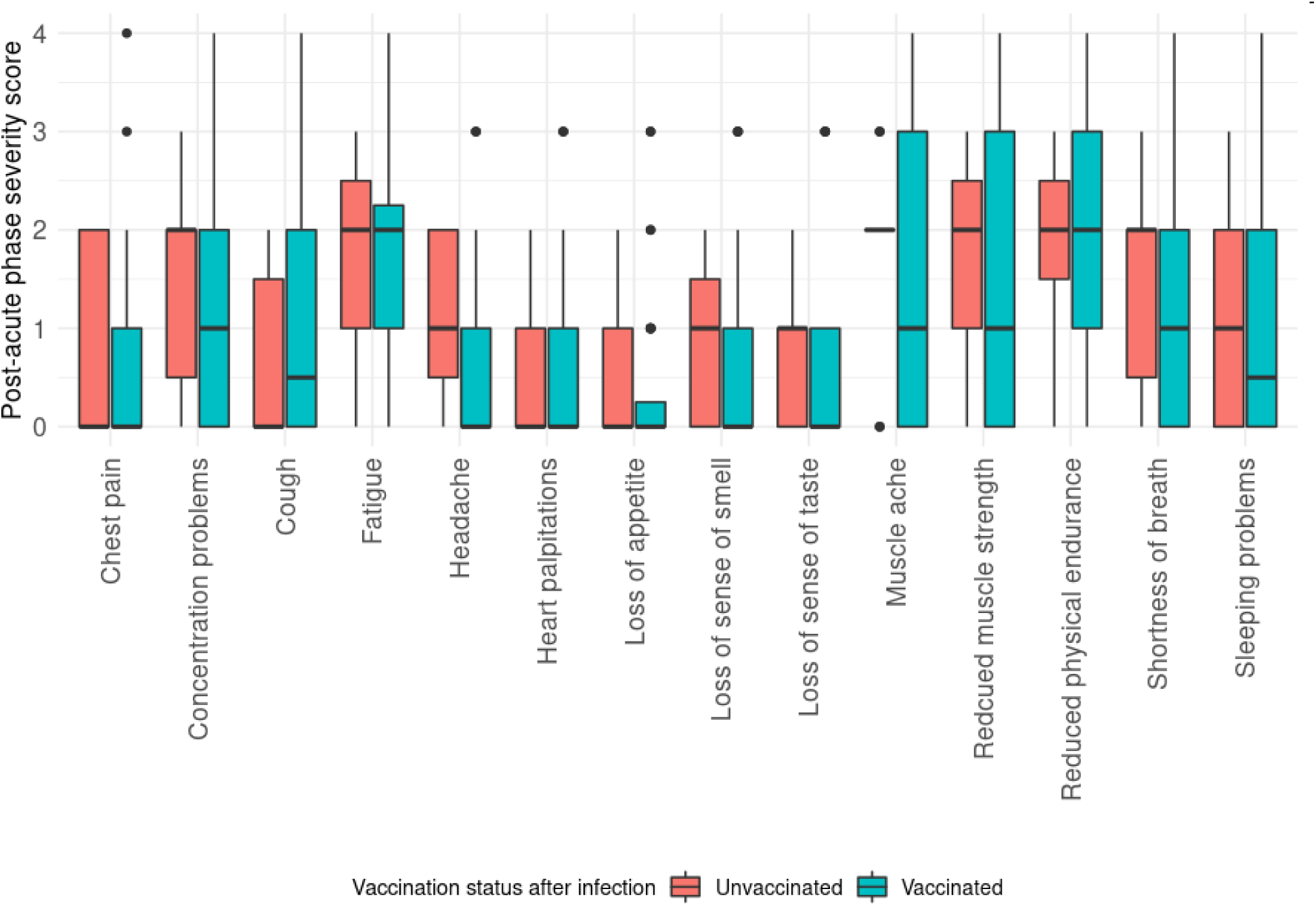
Boxplots showing post-acute phase severity scores for all 14 symptoms and PCC patients (n = 47) stratified by vaccination status after infection. Symptom severity scores range from 0 (did not experience the symptom) to 4 (very severe).

**Supplementary Table S1:** Description of variables.

| Variable | Type | Description |
| --- | --- | --- |
| Change in symptom severity score | Numerical | Main outcome variable. The change in severity score was calculated by subtracting the severity score of a symptom during the post-acute phase from the severity score of a symptom during the acute phase. A positive change in severity score therefore indicates an improvement of symptom severity compared to the acute phase. For both the acute and post-acute phase, symptoms were ranked on a scale of 0 to 4, with 0= absent, 1=mild, 2 = moderate, 3= severe, and 4= very severe. |
| Vaccination status after infection | Binary | Main predictor of interest. This variable indicates whether a PCC patient was vaccinated (yes / no) with either 1 or 2 doses after their SARS-CoV-2 infection. |
| Body Mass Index (BMI) | Numerical | BMI in kg/m<sup>2</sup> |
| Age | Numerical | Age in years |
| Sex | Binary | Binary variable. Male, Female. |
| Acute phase severity score | Numerical | Self-reported severity score for symptom severity during the acute phase of the SARS-CoV-2 infection. Symptoms were ranked on a scale of 0 to 4, with 0= absent, 1=mild, 2 = moderate, 3= severe, and 4= very severe. For each multivariate linear model, this score is specific for the symptom under consideration. |
| Duration of illness | Numerical | Number of weeks between onset of disease date and date of interview. |
| Pre-pandemic health score | Numerical | A self-reported health score of pre-pandemic health (range 0 – 100) using the validated EQ-VAS scale, 0=worst health imaginable, 100=best health imaginable. |
| Smoker | Binary | Binary variable indicating whether a patient smoked cigarettes before the start of the pandemic. |
| Alcohol | Binary | Binary variable indicating whether a patient drank alcohol before the start of the pandemic. |
| Hospitalized | Binary | Binary variable indicating whether a patient was hospitalized for their SARS-CoV-2 infection during the acute phase of infection. |
| Comorbidity | Binary | Variable indicating whether a person suffers from either: Post-acute respiratory problems (including asthma, COPD, TB), post-acute cardiovascular disease, diabetes, severe kidney disease (including dialysis or kidney transplantation), HIV, severe liver disease, being immunocompromised, cancer, depression or anxiety, stroke, rheumatism, arthritis, or memory loss due to a neurological condition or dementia. |

**Supplementary Table S2:** Sensitivity analyses describing the association between duration of illness, post-infection COVID-19 vaccination, and symptom severity change of PCC patients on Bonaire, Caribbean Netherlands.

| Description | Symptom | Adjusting for | Beta Vaccinated (95% CI) | P value |
| --- | --- | --- | --- | --- |
| Original model (see **Supplementary Table 4**) | Heart palpitations | Acute phase severity score, Duration of illness | 0.60 (0.18 – 1.02) | **0.01** |
|  | Shortness of breath | Acute phase severity score, Duration of illness | 0.63 (-0.25 - 1.52) | 0.15 |
| Original model without duration of illness | Heart palpitations | Acute phase severity score | 0.37 (-0.02 – 0.75) | 0.06 |
|  | Shortness of breath | Acute phase severity score | 0.19 (-0.6 – 0.98) | 0.63 |
| New models not including duration of illness in variable selection procedure | Heart palpitations | Acute phase severity score, BMI | 0.30 (-0.08– 0.68) | 0.11 |
|  | Shortness of breath | Acute phase severity score, BMI, Smoker | 0.0 (-0.76 - 0.76) | 1 |
| New models adjusting for duration of illness | Heart palpitations | Acute phase severity score, BMI , Duration of illness | 0.51 (0.09 – 0.94) | **0.02** |
|  | Shortness of breath | Acute phase severity score, BMI, Smoker , Duration of illness | 0.36 (-0.51 – 1.24) | 0.41 |

**Supplementary Table S3:**
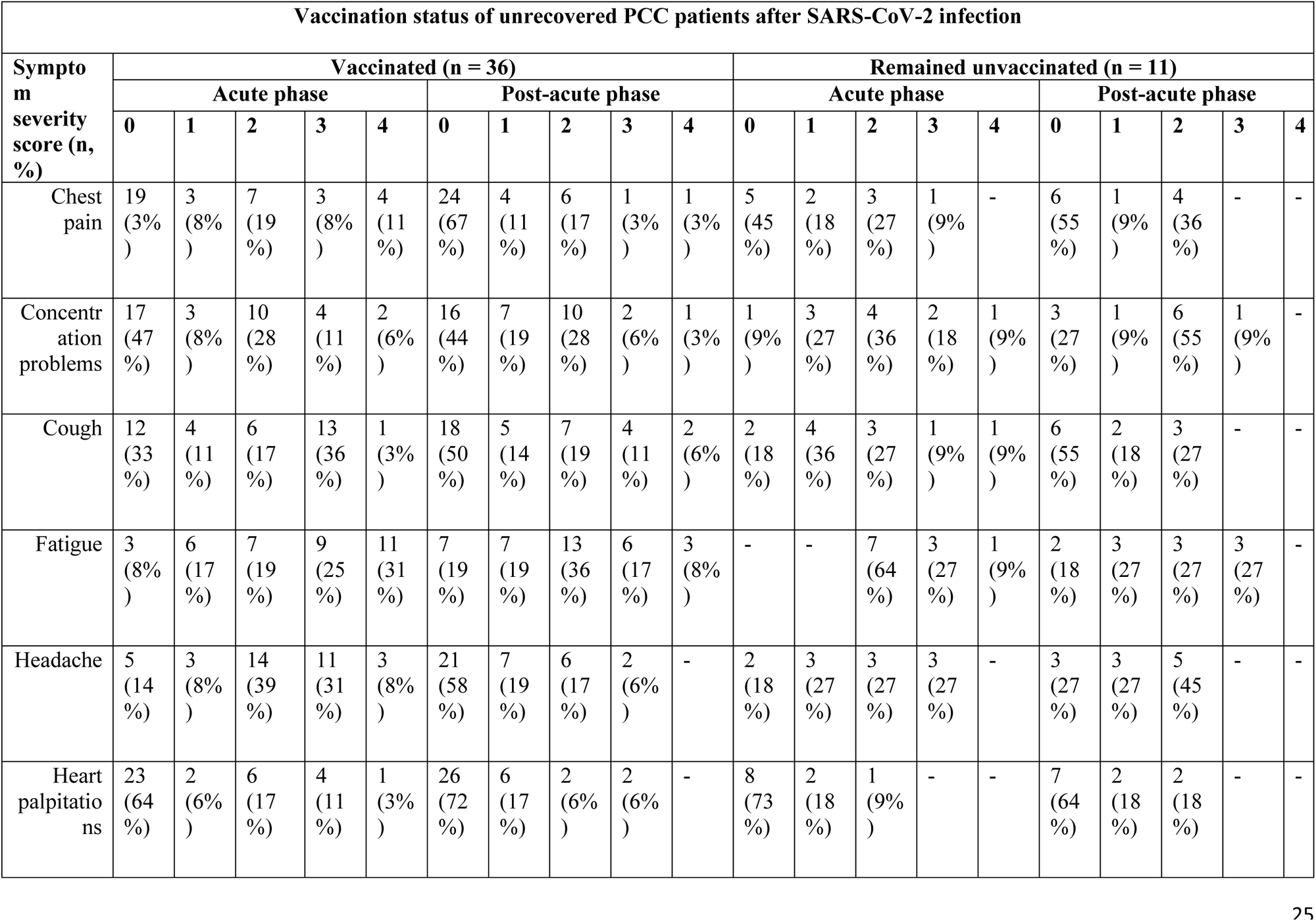
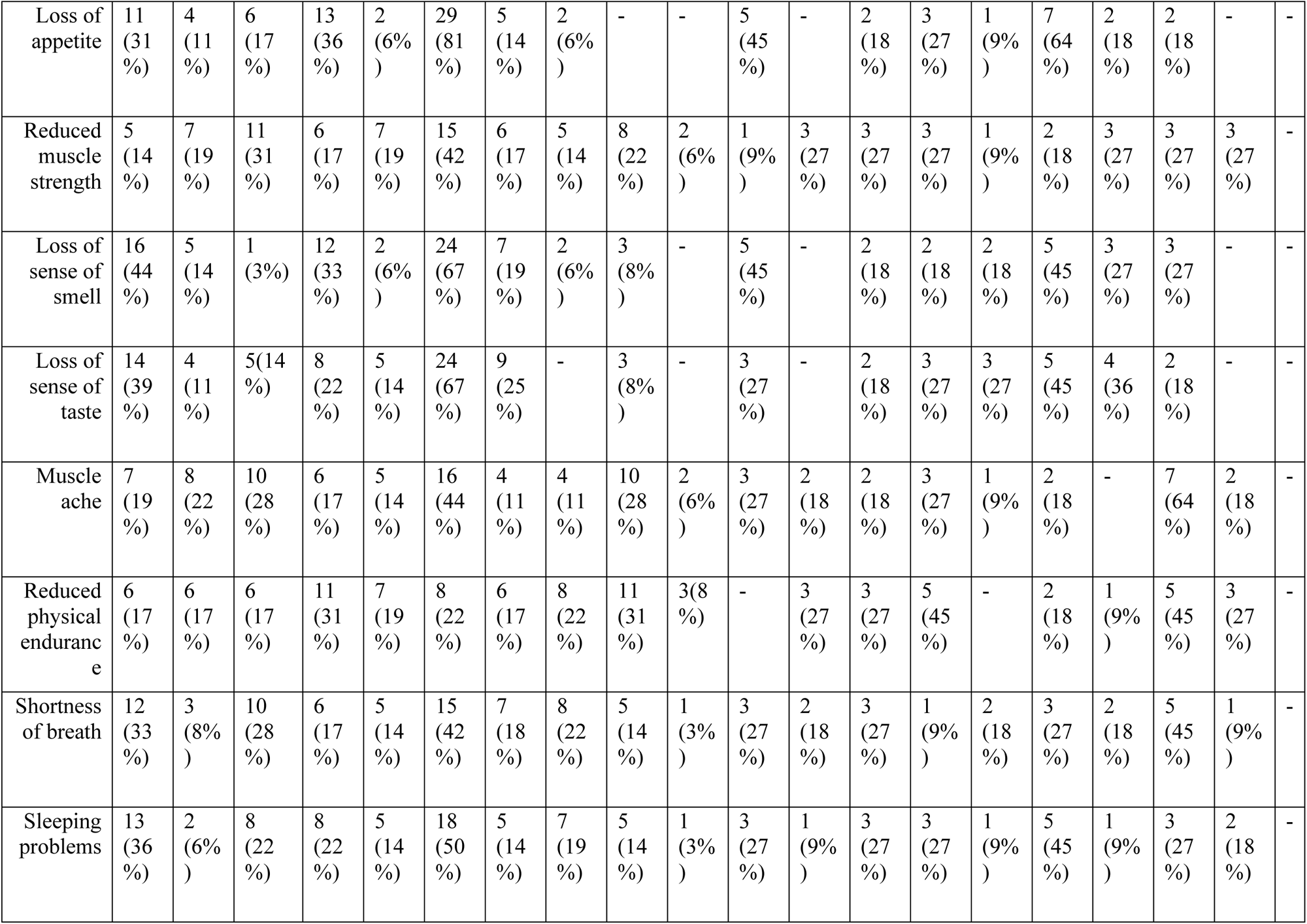
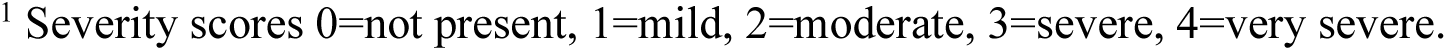
Self-reported severity of COVID-19 symptoms among unrecovered PCC patients, during the acute and post-acute phase of infection, by post-infection COVID-19 vaccination status^1^.

**Supplementary Table S4:** Unadjusted models for the association between post-infection COVID-19 vaccination and symptom severity change of PCC patients on Bonaire, Caribbean Netherlands^1^.

| Symptom | Beta Vaccination Status (95% CI) | P value | AIC<sup>2</sup> |
| --- | --- | --- | --- |
| Chest pain | 0.35 (-0.38- 1.07) | 0.34 | 140.96 |
| Concentration problems | -0.29 (-0.96 -0.38) | 0.39 | 133.79 |
| Cough | -0.26 (-1.26 – 0.73) | 0.60 | 171.21 |
| Fatigue | -0.04 (-0.99 - 0.91) | 0.93 | 166.59 |
| Headache | 0.96 (0.10 - 1.82) | **0.03** | 157.75 |
| Heart palpitations | 0.57 (0.06 - 1.08) | **0.03** | 108.13 |
| Loss of appetite | 0.33 (-0.59 - 1.26) | 0.47 | 164.39 |
| Reduced muscle strength | 0.39 (-0.44 - 1.21) | 0.35 | 153.37 |
| Loss of sense of smell | 0.04 (-0.78 - 0.87) | 0.92 | 153.85 |
| Loss of sense of taste | -0.43 (-1.37- 0.50) | 0.36 | 165.70 |
| Muscle ache | 0.54 (-0.51 -1.58) | 0.31 | 175.70 |
| Reduced physical endurance | -0.03 (-1.08 - 1.02) | 0.95 | 176.05 |
| Shortness of breath | 0.16 (-0.96 - 1.29) | 0.77 | 182.45 |
| Sleeping problems | 0.03 (-0.81 -0.87) | 0.94 | 155.72 |

<sup>1</sup> The reference category is being unvaccinated before the SARS-CoV-2 infection.
<sup>2</sup> Akaike information criterion.

**Supplementary Table S5:**
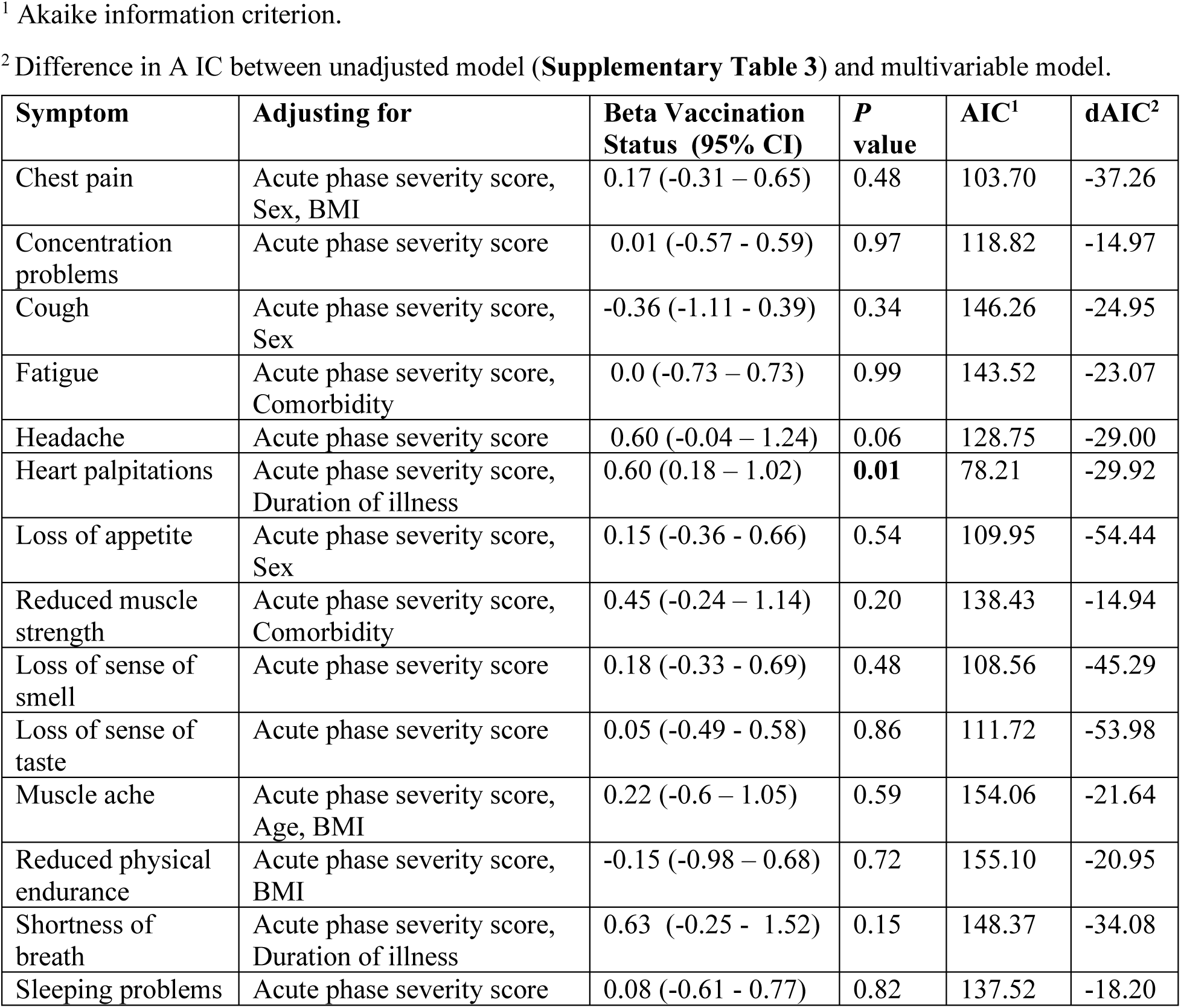
Fully adjusted models for the association between post-infection COVID-19 vaccination and symptom severity change of PCC patients on Bonaire, Caribbean Netherlands.

